# Brazil Health Care System preparation against COVID-19

**DOI:** 10.1101/2020.05.09.20096719

**Authors:** Lincoln Luís Silva, Amanda de Carvalho Dutra, Pedro Henrique Iora, Guilherme Luiz Rodrigues Ramajo, Gabriel Antônio Fernandes Messias, Iago Amado Peres Gualda, Joao Felipe Hermann Costa Scheidt, Pedro Vasconcelos Maia do Amaral, Catherine Staton, Thiago Augusto Hernandes Rocha, Luciano de Andrade, João Ricardo Nickenig Vissoci

## Abstract

**Background:** The coronavirus disease outbreak from 2019 (COVID-19) is associated with a severe acute respiratory syndrome coronavirus 2 (SARS-CoV-2), a highly contagious virus that claimed thousands of lives around the world and disrupted the health system in many countries. The assessment of emergency capacity in every country is a necessary part of the COVID-19 response efforts. Thus, it is extremely recommended to evaluate the health care system to prepare the country to tackle COVID-19 challenges.

**Methods and Findings:** A retrospective and ecological study was performed with data retrieved from the public national healthcare database (DATASUS). Numbers of intensive care unit and infirmary beds, general or intensivists physicians, nurses, nursing technicians, and ventilators from each Regional Health Unity were extracted, and the beds per health professionals and ventilators per population rates were assessed. The accessibility to health services was also performed using a spatial overlay approach to verify regions that lack assistance. It was found that Brazil lacks equity, integrity, and may struggle to assist with high complexity for the COVID-19 patients in many regions of the country.

**Conclusions:** Brazil’s health system is insufficient to tackle the COVID-19 in some regions of the country where the coronavirus may be responsible for high rates of morbidity and mortality.

## INTRODUCTION

The coronavirus disease 2019 (COVID-19) is associated to the novel severe acute respiratory syndrome coronavirus-2 (SARS-Cov-2) identified in December 2019 (1). As of May 4th, 2020 COVID-19 has globally infected 3,759,967 people resulting in 259,474 deaths (2). The World Health Organization (WHO) declared COVID-19 a public health emergency of international concern (PHEIC) by the end of January 2020 under the International Health Regulations (3). Few weeks after the PHEIC declaration, the COVID-19 outbreak was declared to be a pandemic, drawing attention worldwide (4).

The pandemic led to the adoption of several non-pharmacological interventions ranging from social distancing guidelines to national-level lockdowns by different countries (5). These stringent interventions have severely impacted the way of living of many people, and disrupted the already precarious health system in many countries (6). In response to the COVID-19 pandemic, several countries undertook analysis for the necessary health system strengthening efforts. According to studies dedicated to characterize the clinical evolution of the disease 12% of the cases demanded ICU in Italy (7). In the U.S the percentage of patients needing ventilator support was even higher, reaching up to 12,2% (8).

The response effort to tackle the COVID-19 requires a strong organization of the emergency network (9). The lack of beds, iniquities in distribution of hospitals, and an inadequate availability of ventilators could hamper the actions aiming to decrease the negative consequences of the COVID-19 (7). Unfortunately, usually the distribution of the health resources within the countries are characterized by inequities (10). Due to the COVID-19 consequences, the scenario faced by low and medium income countries is even more staggering (7). The historic challenges regarding an insufficient number of health professionals, iniquities in distribution of human resources (11), low accessibility to emergency care services (ECS) (12) and economic issues creates additional pressures to be addressed, aiming is to achieve an adequate COVID-19 response.

As the COVID-19 spreads around the world, the hospital systems lack measures against the virus (13), and many countries are experiencing shortages of hospital supplies (14). For example, as of March 11th, in Italy, where there were 12,462 cases of COVID-19 and 827 deaths, 1,028 of 5,200 beds in intensive care units (ICU) are occupied. A few days later, there were no more ICU beds available (15). In the United States of America, it is estimated that the disease will stress bed capacity, equipment, and health care personnel as never seen before (16). The Brazilian case is not an exception (12). In order to reduce the burden, the hospital administrators, governments, policy-makers, and researchers must prepare for a surge in the Healthcare System (17).

On May 7th there were 149,988 confirmed cases and 9,600 deaths in Brazil (18). However, this number is under-reported and the real number is estimated to be nine times greater according to some simulations (19). To further the concern, The Imperial College estimates that up to 1 million of people will fall ill due to COVID-19 in Brazil (20).

During the last decade Brazil is struggling to increase the funding of the public unified system (SUS). Despite the efforts performed in 2016 the Constitution Amendment 95 (EC 95 acronym in Portuguese) reduced the budget of the Ministry of Health by almost seven billions Reais by year (21). Before the EC95 the Brazilian Health System was already underfunded (22). Two years after the EC95 the consequences regarding the lack of funding were aggravated by the COVID-19 pandemic. Additionally, Brazil is also facing a political crisis contributing to divergences between the administrative levels in the country.

The consequences of all these elements combined could hamper the response actions to tackle the COVID-19. The availability of information during a crisis is essential to support the decision-making process based on evidence. Taking this point into consideration the present work addresses critical aspects regarding the organization of the emergency network system in Brazil, jointly with the spatial expansion of COVID-19 cases within the country, and to highlight where the efforts currently performed in Brazil were capable of coping with the lack of access to emergency care needed to cope COVID-19 consequences.

## METHODS

### Study design and local

The present paper is an ecological, observational and cross-sectional study using spatial analysis approach. The data sources is based on secondary data from the Unified Health System (SUS) (23). To fulfill the defined objective the adequacy parameters in terms of human resources, health care structure and accessibility to emergency care services (ECS) were analyzed in comparison with the reported incidence of COVID-19.

According to data from the Brazilian Institute of Geography and Statistics (IBGE), Brazil is located in South America with a territorial area of 8.510.820,623 km2, and a total of 210,147,125 inhabitants, with Human Development Index (HDI) of 0.51 with diversified values for the municipalities ranging from 0.41 to 0.86 (figure 1) (24). For the assessment of methodological quality, we followed the Guideline Strengthening the Reporting of Observational Studies in Epidemiology (STROBE) (25).

**Figure 1.**
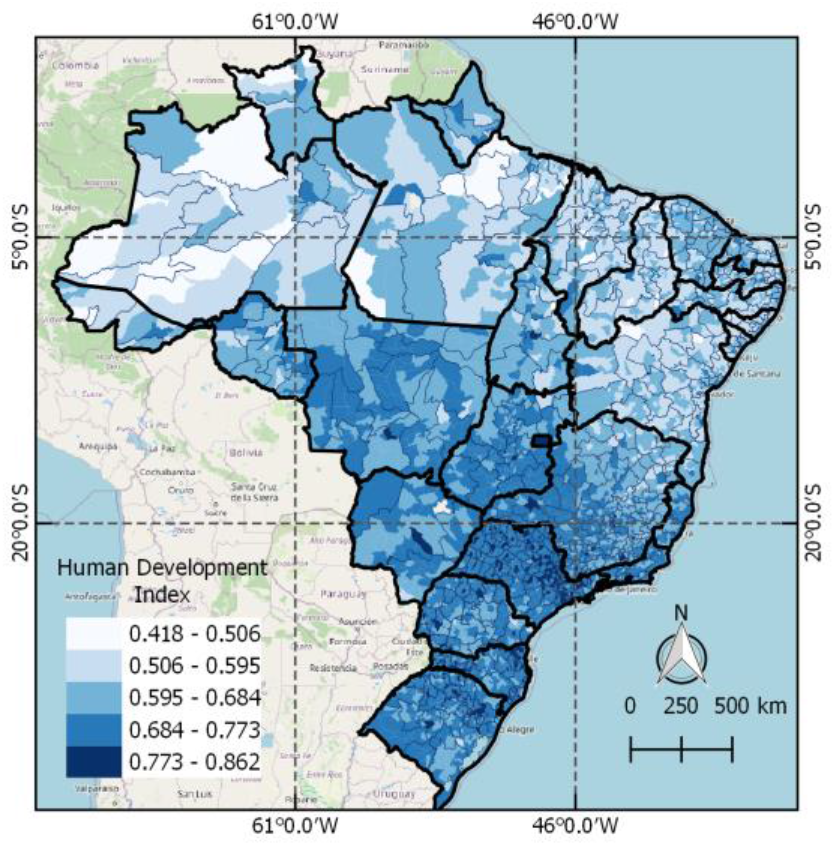
Location of Brazil and distribution of Human Development Index.

### Data sources

To characterize the Brazilian ECS network, three sources were used: National Register of Health Facilities (CNES acronym in Portuguese), population data from the IBGE, and COVID-19 cases reported by Secretariat of Surveillance of the Ministry of Health (18, 26). Data regarding hospitals, professionals (nurses, nursing technicians, nursing assistants, doctors and physiotherapists), and equipment (ventilators, ICU and infirmary beds) were obtained from the CNES website using R through the microdataus package (27). Only adult and pediatric ICUs of type I, II and III were collected as intended for patients with COVID-19 according to ordinance 414 of the Ministry of Health. (28). The population data and thematic maps were fetched from the IBGE (24).

The match between the number of health professionals and the recommended suitability parameters were compared using the guidelines from the National Health Surveillance Agency (ANVISA) Resolution of the Collegiate Board of Directors (RDC). The ANVISA RDC number 7 provides the minimum requirements for the operation of Intensive Care Units, in which ten ICU beds are required for each one intensive care physician and one physiotherapist, one intensive care nurse for each eight beds, and two nursing assistants for each bed (29). The RDC also has recommends 1 ventilator per 2 beds (29).

The building of thematic maps was carried out by grouping the municipalities by Health Regions Unity (H.R.) using software QGIS 3.0. The (H.R.) is a continuous geographic space constituted by a group of bordering municipalities delimited by cultural, economic and social identities, created by the Ministry of Health in order to mitigate the disparities in the country (30).

### Spatial distribution of COVID-19 cases and the lack of emergency care

To identify regions with a high incidence of COVID-19, simultaneously presenting a lack of emergency network was used as a spatial overlay approach. The first step comprised the development of an emergency infrastructure index (EII). The EII was obtained computing the number of beds registered, by the ratio of professionals and equipment according to the last competence of February 2020 from CNES.

To evaluate the geographical accessibility to emergency care service care was used the two-step floating catchment area (2SFCA) technique. With this approach, it was possible to assess the accessibility to emergency care services by the interaction of two geographic characteristics: (a) the volume of available hospital beds weighted by population within 2 hours of travel distance, and (b) the proximity of hospitals within a 2 hours displacement from each municipality (11). The 2SFCA method generated two accessibility indexes for each municipality in Brazil, one regarding the network available in February 2020, and a secondary one highlighting where the COVID-19 new exclusive beds increased the access to emergency services. Both indexes created the conditions to identify regions with a lack of access to emergency care, as well as the regions being benefited by the expansion of the ICU beds dedicated to the CODVID-19 response. To highlight regions with a high incidence of COVID-19 and a lack of emergency structure, an overlap analysis was conducted to select the municipalities concurrently, showing a pattern of high incidence, jointly with a lack of access to emergency services.

Once the EII was computed, and the municipalities with high incidence within regions with low access to emergency care services care were identified, a Getis-Ord-Gi analysis was performed. Thus, it was possible to point out three spatial clusters: (1) emergency care services accessibility on February 20; (2) municipalities with low access to emergency care services and high COVID-19 incidence, (3) accessibility to ICU beds exclusively dedicated to COVID-19 response in March 2020.

### Ethics

In accordance with the Resolution No. 510/16 of the National Health Council and considering that we used secondary sources which are available in governmental and online databases, the dispensation of the consent form was requested to the Ethics Committee.

## RESULTS

All of 5,570 municipalities are distributed in 5 regions and 438 Health Regions Unity in Brazil. Besides that, Brazil has a total of 35,682 ICU beds, 426,388 hospital beds, 65,411 ventilators, 18,716 intensivists, 564,529 general physicians, 2,768 intensive care nurses, 263,315 nurses, 710,846 nursing technician and 83,040 physiotherapist registered in the database (February, 2020). In terms of professionals and beds per 10,000 inhabitants, the southern region had the greatest rates for ICU beds, ventilators, physicians, nurses and technicians, while the north region had the lowest ones (Table 1). The figure 2 shows the rates of beds and professionals per Health Regions. The ICU beds per intensivist varied from zero to 53, zero to 156.5 per intensive care nurse, zero to 0.21 per technician, and zero to 2 per physiotherapist. In addition, hospital beds per physician varied from 0.10 to 7.14, 0.52 to 11.26 per nurses, 0.19 to 2.58 per technician, and 2.72 to 13.37 per physiotherapist.

**Table 1.** Rates of different types of bed and professionals per population in Brazil.

|  | ICU beds | Infirmiry beds | Physicians | Nurses | Technicians | Ventilators |
| --- | --- | --- | --- | --- | --- | --- |
| South | 1.70 | 24.33 | 31.72 | 13.21 | 35.46 | 3.13 |
| Southern | 2.11 | 19.46 | 34.70 | 14.14 | 38.57 | 3.82 |
| Midwest | 2.08 | 22.64 | 23.85 | 9.90 | 33.17 | 3.78 |
| Northeast | 1.19 | 20.00 | 17.60 | 11.62 | 27.39 | 2.19 |
| North | 0.95 | 16.47 | 12.74 | 8.82 | 28.87 | 1.91 |
| Brazil | 1.70 | 20.29 | 26.86 | 12.53 | 33.82 | 3.11 |

\*According to the IBGE Cidades, it is estimated 210,147,125 inhabitants in Brazil.

**Figure 2.**
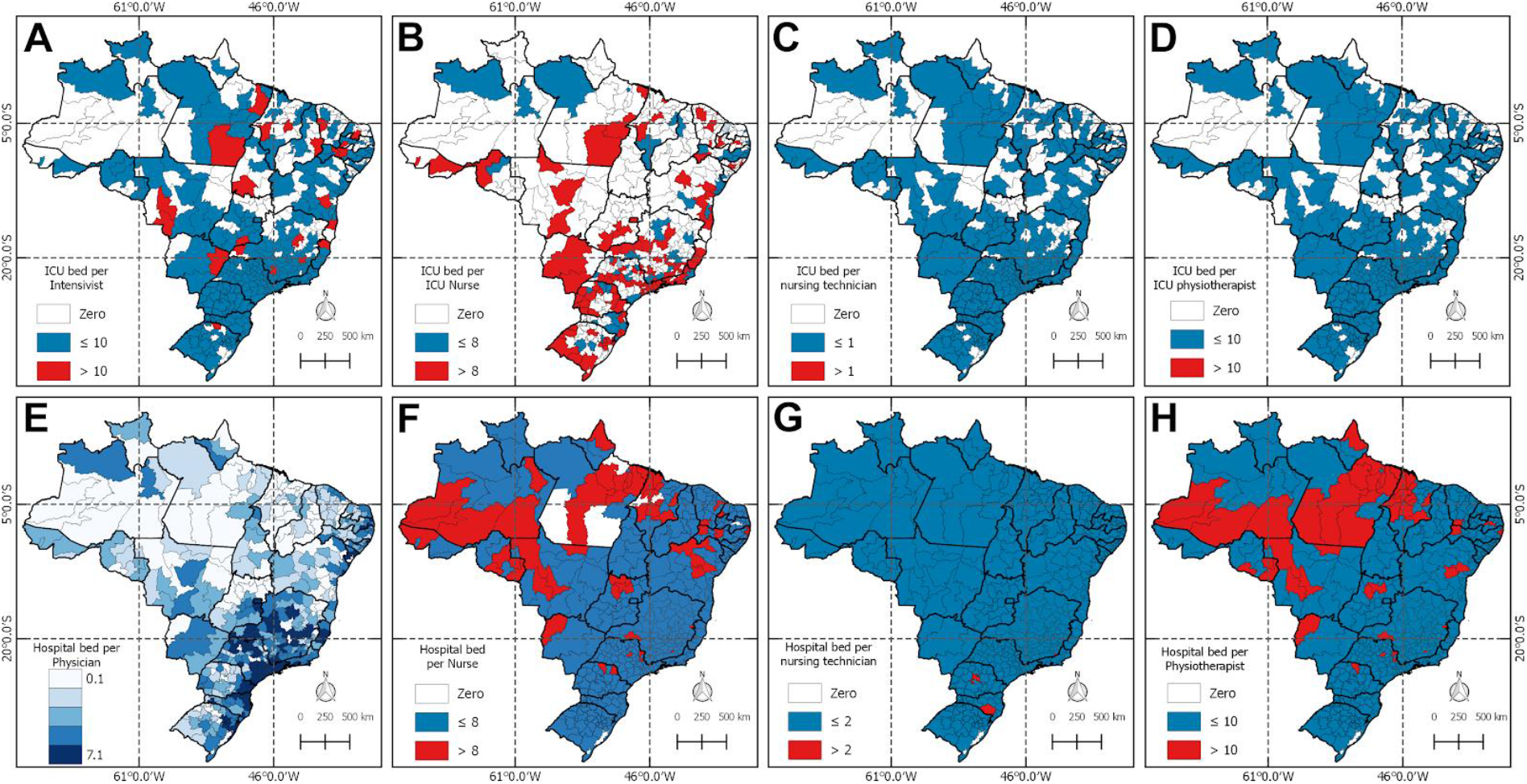
Regions of Brazil where the conditions of beds, equipment and professionals are demonstrated according to the Resolution of the Collegiate Board of Directors (RDC).

Figure 2 shows the distribution of professionals, beds and ventilators throughout the territory, and classifies this distribution according to RDC number 7. In A, most H.R. are in accordance with the RDC, which determines up to 10 intensivists for each ICU bed. However, 132 H.R. do not have ICU beds and / or have no intensivist. In B, only 54 H.R. are working according to the recommended amount of 1 critical care nurse for each 8 ICU beds. C and D show that 322 H.R. are working correctly with the capacity of 1 nursing assistant and 1 physiotherapist for each 2 and 10 beds, respectively. In the second row of figure 2, F, G and H show that there are nurses, nursing technicians and physiotherapists working above the limit of 1 nurse per 8 infirmary bed, 1 nursing technician for each 2 infirmary bed and 1 physiotherapist per 10 infirmary bed.

COVID-19 has shown a fast growth rate in Brazil as shown in figure 3A. In addition, the growth rate of confirmed deaths shows a very unequal pattern in some regions. Likely due to different state-level isolation policies, the states of São Paulo, Rio de Janeiro, Ceará and Amazona have shown a much faster growth rate than the rest of the country (Figure 3B). Nonetheless, the country as a whole seems to be still far from its peak number of new deaths by COVID-19.

**Figure 3.**
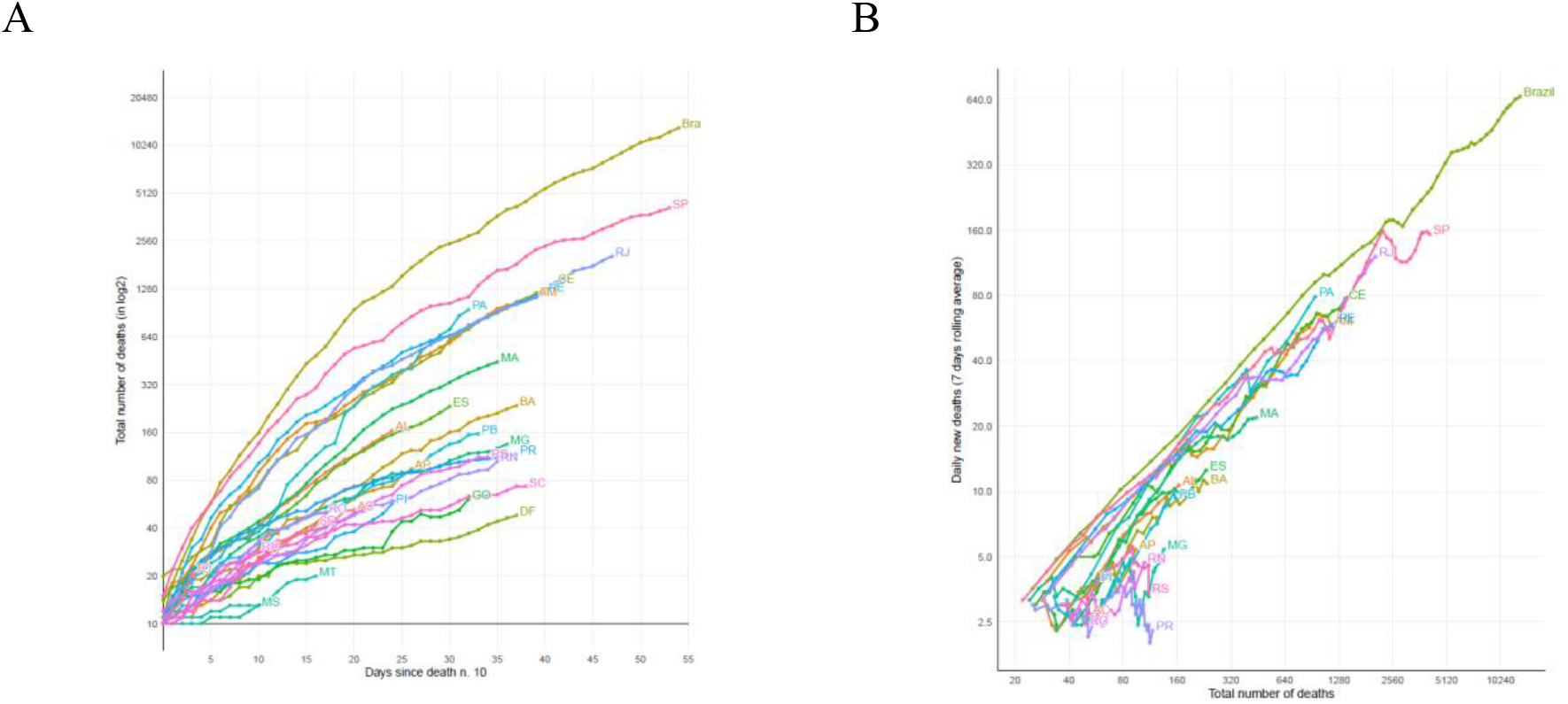
A. Total number of deaths in selected countries by days since 10th death. B. Total number of confirmed deaths and daily new deaths by COVID-19 by selected countries and Brazilian states (on 07-May-2020). Source: Data from the European Centre for Disease Prevention and Control (ECDC) and Brazilian Ministry of Health.

Figure 4 shows the distribution of 149,988 cases of COVID-19 on May 7th and location of 12,428 new adult and pediatric ICU beds available on April 27th. Most cases and new ICU beds are located in the southeast region, while the north region received few beds in comparison with other regions. Figure 5 presents the distribution of ICU beds per ventilator and shows that in the north there are no ventilators available in some H.R.

**Figure 4.**
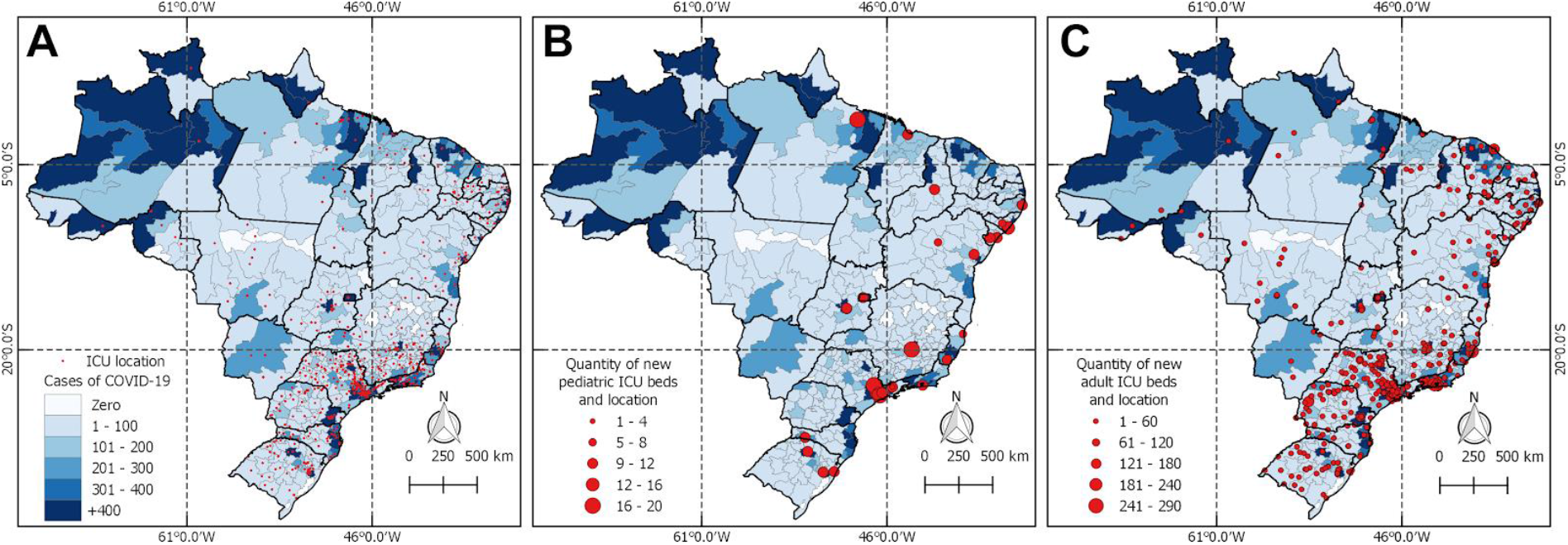
A) Location of intensive care units (ICU) and the number of cases of COVID-19 in Brazil as of April 27th. B) Distribution of new pediatric ICU beds. C) Distribution of new adult ICU beds.

**Figure 5.**
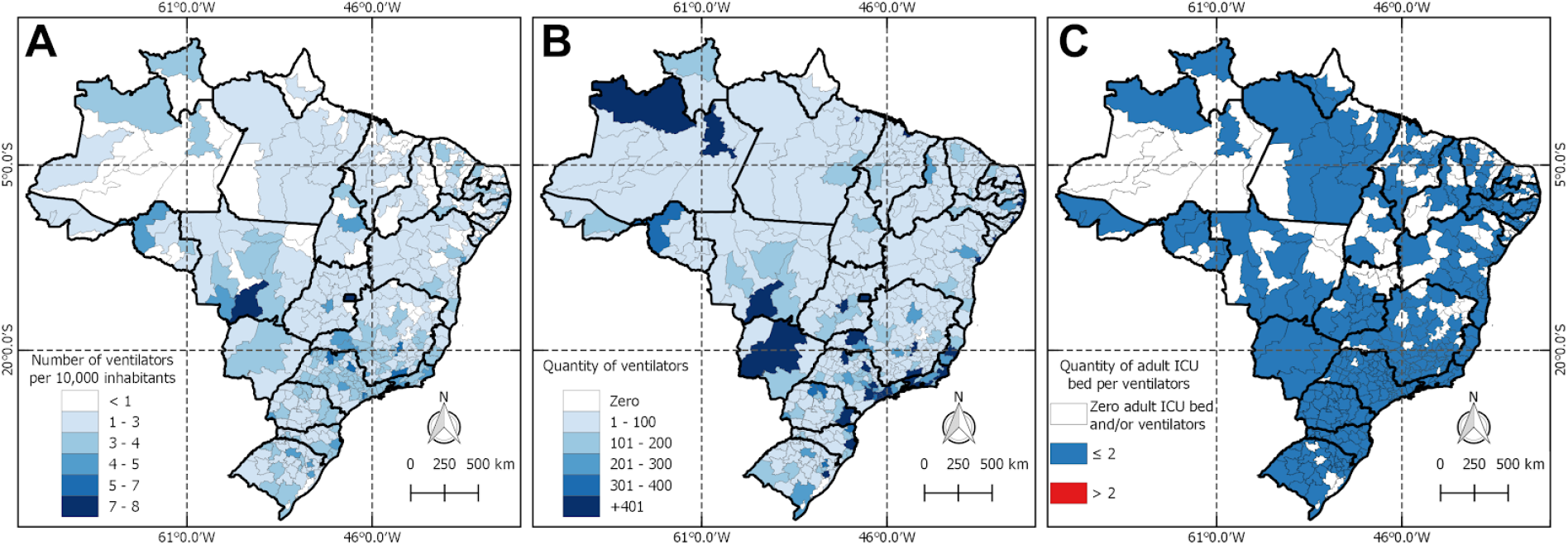
A) Distribution of ventilators in Brazil for every 10,000 inhabitants. B) Number of ventilators in each Health Region. C) Ventilators per bed according to Collegiate Board Resolution.

The panel A in the figure 6 depicts the COVID-19 incidence by Brazilian municipality up to 04/29/2020. Is possible to observe that all states and regions currently are presenting COVID-19 cases with a highlight to the states of Amazonas, Amapá, Espiríto Santo and Santa Caterina where it is already noted areas with hot colors. The hot colors indicate higher levels of incidence. In terms of deaths all state capitals of the North, Northeast and southeast regions are presenting high levels of mortality when compared with the rest of the country.

**Figure 6.**
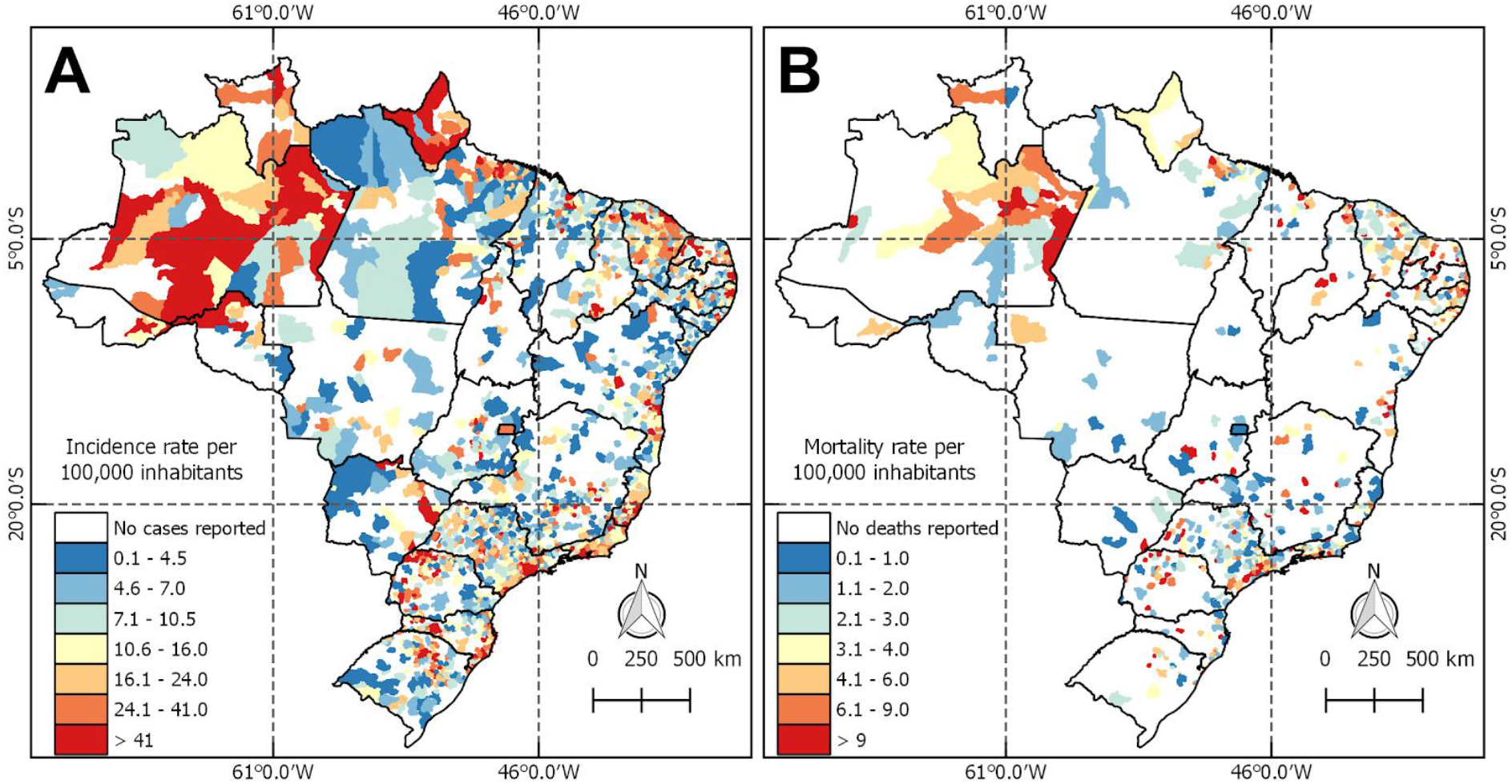
COVID-19 incidence and mortality rate in Brazil.

The figure 7 presents three maps characterizing the Brazilian situation in terms of emergency services and the COVID-19 incidence. The panel A exhibits the accessibility index to ICU beds by population. The map highlights a higher accessibility index close to the state capitals of the Brazilian states. The map B emphasizes the municipalities presenting a COVID-19 incidence higher than the national average of 16.49, simultaneously with an accessibility index lower than the national mean 21 per 100,000 inhabitants Thus, every municipality in the map B is facing challenges in terms of ECS and a high COVID-19 incidence for the Brazilian standards. The map C presents the accessibility index of the new beds created exclusively to offer intensive care to COVID-19 patients. Few beds with this specific purpose were open in the states of the North and Midwest region of the country.

**Figure 7.**
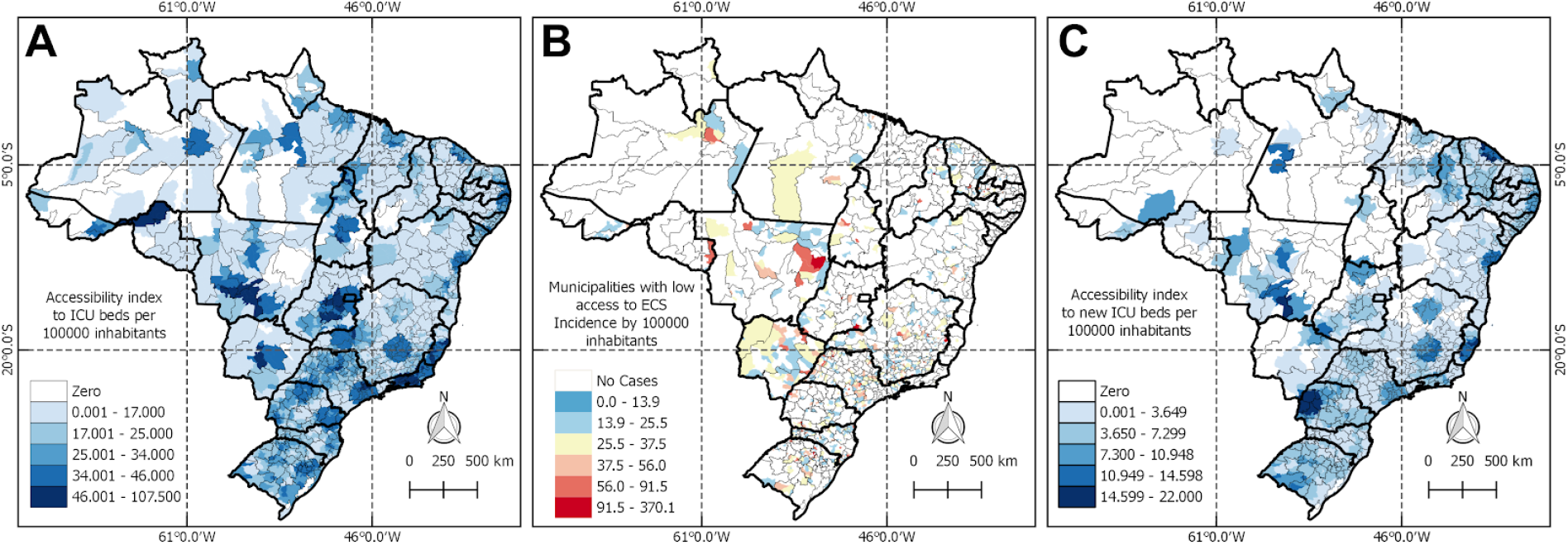
Scenario of emergency care services network to tackle COVID-19, 2020, Brazil. The panel A shows the accessibility index on February 2020. B presents the high incidence of the disease and the lack of emergency care services. C exhibits the accessibility index of new beds created to tackle the COVID-19.

The figure 8 shows the result of spatial clustering analysis to identify trends in access, as well as in the COVID-19 incidence. Panel A exhibits a hot spot covering the Southeast, Midwest, and South regions of the country. The states covered by the red layer presents the spatially significant group of municipalities with higher levels of access to ICU beds by population. On the opposite side, the blue layer highlights the regions facing geographical barriers to grant access to ICU beds to the population. To build the map B, the same approach was used, but this time only applied to the municipalities with high COVID-19 incidence and a low index of accessibility to ICU beds. The red color characterized a group of municipalities in the South and Midwest regions. Despite the higher availability of beds in these regions, it was possible to observe a statistically significant group of municipalities within these regions with barriers to access ICU beds. Map C illustrates the cluster of accessibility regarding the ICU beds created to tackle the COVID-19. The lack of overlay between the red color of maps B and C is pointing out a mismatch of the response efforts dedicated to addressing the COVID-19 challenge. The regions in map B characterized as hotspots were considered cold spots regarding the creation of ICU beds dedicated to COVID-19. The result suggests that the use of scarce resources needed to put in order ICU beds are not being directed to municipalities lacking access to emergency care services, despite their high levels of COVID-19 incidence.

**Figure 8.**
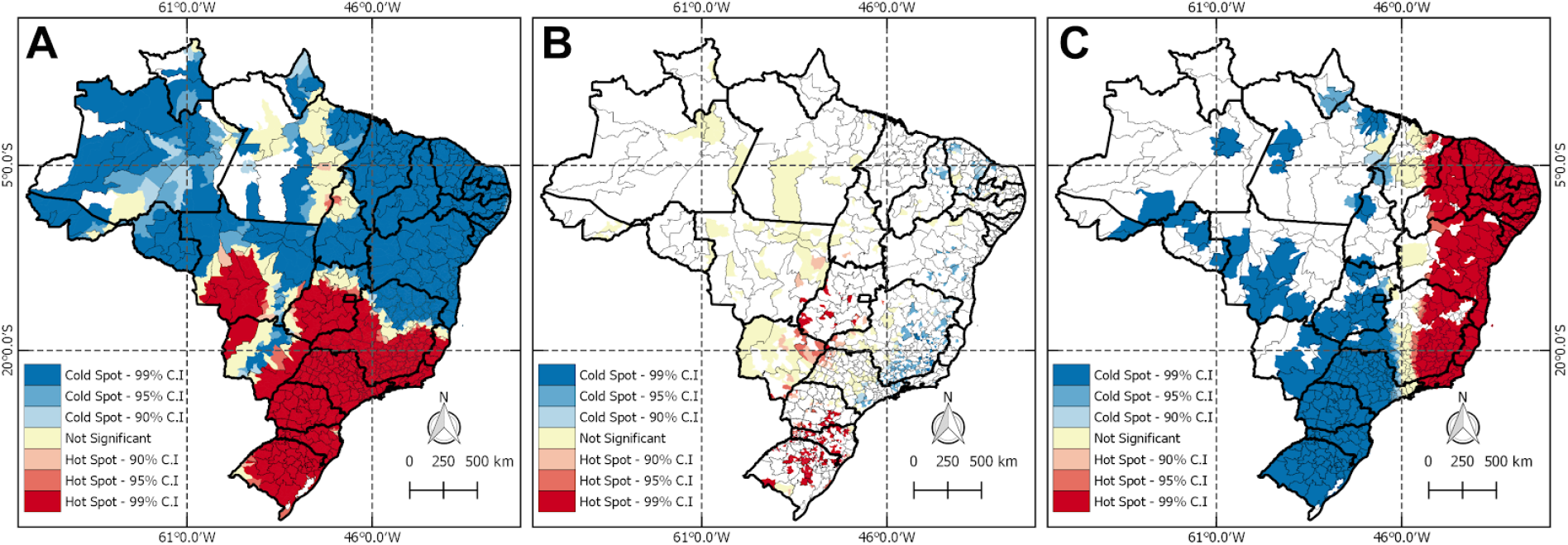
Spatial clusters of access to ECS dedicated to tackle COVID-19, 2020, Brazil. The panel A shows the accessibility index on February 2020. B presents the high incidence of the disease and the lack of emergency care services. C exhibits the accessibility index of new beds created to tackle the COVID-19.

## DISCUSSION

The ongoing COVID-19 pandemic has caused nearly 4 million confirmed cases and claimed over 278,756 lives worldwide as of 9th May 2020 (2). It is noteworthy to mention that the COVID-19 outbreak is a challenge to the health systems worldwide (31), and although the outcome for the crisis caused by this disease is uncertain, SARS-CoV-2 will overwhelm the health care infrastructure for months (32). In this study, the Brazilian health system was evaluated in order to verify its capacity to tackle the COVID-19 challenge.

According to the WHO, it is recommended one doctor and one nurse per 1,000 inhabitants as a parameter of health care for the population (33). To strengthen the WHO recommendations, the Brazilian Health Governments has established in the Resolution of the Collegiate Board of Directors (RDC) the quantities of ICU and infirmary beds per intensivists, general physician, nurses, nursing technician and physiotherapists (29). Although Table 1 shows that in Brazil there is sufficient number of physicians in the country, the figure 2 shows that these professionals are not evenly distributed to accomplish the WHO recommendations. In addition, the number of nurses do not meet the criteria in the north and Midwest. To illustrate the problem, figure 2 shows that Brazil has desert zones of ICU assistance and regions where these professionals have to take care of beds far beyond the quantities stipulated by the RDC. Bahtt et al., 2017 verified that professionals in critical care that were caring for more patients per shift were more likely to experience burnout (34). Halpern et al., 2017 informed that intensivists are also in shortage in the United States of America and this situation may be attributed to burnout (35). Therefore, the combat against the COVID-19 may be a difficult task in these regions, since providing access and affordable care for the large urban populations is already a challenge for many countries (36).

Experience from Lombardia has shown that 9% of patients were admitted in the ICU treatment, whereas this number varied from 5 to 32% in some cities in China (37). On the other hand, in Brazil there’s no available large data of ICU patients at the moment, and supposing that those numbers might appear in the country as well, only 4 out of 438 Health Regionals could manage this amount of patients.

In terms of nursing care in ICU accessibility, figure 2 shows that there are large regions of assistential voids, probably because there are low amounts of ICU nurses in Brazil (38). Besides that, it’s possible to visualize that there are regions where ICU and generalist nurses are responsible for more than eight ICU and infirmary beds, which may represent a risk of unfavorable outcome for the COVID-19 treatment since that high amount of patients per nurse are associated with a range of negative patient outcomes (39, 40).

The pandemic has led to severe shortages of many essential supplies such as ICU beds and ventilators (41). Based on Italy’s numbers that 10 to 25% of hospitalized patients will require ventilation, the Centers for Disease Control and Prevention estimates that in the USA there will be between 1.4 to 31 patients per ventilator this period (30, 41). Brazil, on the other hand, 2.5 to 3.4 million people will require hospitalization, according to The Imperial College of London (20), which represents 4 to 13 patients per ventilator if distributed equally. The numbers may represent a satisfactory amount of equipment, however, figure 5 shows that there’s no ventilators in some H.R. that are potential places to have a high number of deaths.

The most concerning result was obtained through the spatial cluster analysis. As referring in a recent editorial by the Lancet, Brazil seems to be facing a double crisis. The Lancet editorial highlighted that the federal government’s is going against the technical recommendations of the Ministry of Health and WHO. Consequently, early on the facing the pandemic there was a disagreement between the Federal administration, and the states and municipalities. On account of that situation, each administrative level implemented isolated responses to prepare for the the COVID-19 with limited country-level coordination. The spatial clusters analysis highlighted that new beds created to tackle the COVID-19 were not distributed according to the gaps in access in care historically known in Brazil, potentially a result of the lack of central planning and governing in facing the crisis. The hot spot clusters of municipalities with high incidence and lack of access are not overlapping with the hot spot cluster of new beds dedicated to the COVID-19.

From now on, Brazil has several difficulties in treating patients in critical care. This paper shows that there is an insufficient number of ICU beds, ventilators, and a huge lack of professionals in healthcare. Additionally, the allocation of new beds aiming to fight the COVID-19 pandemic did not address areas with known gaps in care and health infra-structure availability, contributes to worsening the situation observed through the other indicators assessed. Developed countries like Italy and the United States demonstrate that COVID-19 can overwhelm the healthcare capacities of well-resourced nations very fast (16,38). Therefore, the SARS-CoV-2 epidemic in middle-income countries, such as Brazil (39), may be devastating. Our findings suggest that strong leadership is needed to coordinate the response efforts against the COVID-19.

The limitations in this work rely on the complex data available. Health data from health information systems, including health-facility records, surveys or vital statistics, may not be representative of the entire population of a country and in some cases may not even be accurate (44). The CNES database presents some limitations well known by the Brazilian scientific community (45). Despite this, the information regarding the availability of COVID-19 beds was published just a month ago, calling attention to the occurrence of efforts aiming to improve the quality of the data available to policymakers.

## Data Availability

All data used is publicly available at http://tabnet.datasus.gov.br/cgi/tabcgi.exe?cnes/cnv/prid02mg.def

## ACKNOWLEDGMENTS

We would like to thank the Coordination for the Improvement of Higher Education Personnel (CAPES).

## AUTHOR CONTRIBUTION

LLS, ACD and PHI collected the data, wrote the paper and performed the illustrations. GLRR, GAFM, IAPG, JFHCS, PVMA cleared the bank, validate the datasets and translated the manuscript. CS, TAHR, LA performed the analysis of accessibility and revised the manuscript. JRNV led the team, designed the study and took the final decision of publishment.

